# LAG3 as an independent TME biomarker in Chinese colorectal cancer: Validation of a lung cancer-derived subtyping signature

**DOI:** 10.64898/2026.08.04.26359661

**Authors:** JinLong Li, JingXian Huang, ZhouHuan Dong, YiXin Huo

**Affiliations:** Beijing Institute of Technology,No. 5, South Street, Zhongguancun, Haidian District, Beijing, China 100081; Pathology Department, First Medical Center, Chinese People’s Liberation Army General Hospital, No. 28, Fuxing Road, Haidian District, Beijing,China 100853; Yantai Institute of Technology, No. 100 Gangcheng East Street, Laishan District, Yantai, Shandong, China 264003

**Keywords:** Colorectal pathology, Tumor microenvironment, Immune pathological biomarker, LAG3, IRF1, CD8A, Clinical molecular pathology;next-generation sequencing

## Abstract

**Objective:** Commercially available next-generation sequencing (NGS) platforms in China routinely adopt a lung cancer-derived tumor microenvironment (TME) subtyping signature from a European cohort to classify colorectal cancer (CRC), yet its diagnostic performance in Chinese CRC patients remains unvalidated. This study aimed to evaluate the subtyping efficiency of the lung cancer TME signature in a Chinese CRC cohort, screen CRC-specific immune mRNA biomarkers for TME subtyping, and explore the clinical utility of IRF1, CD8A and LAG3 for distinguishing immune-enriched (IE) and immune-desert plus fibrotic (D+F) subtypes.

**Methods:** A total of 87 FFPE CRC specimens with complete NGS and clinicopathological data were retrospectively enrolled, including 15 IE subtype and 72 D+F subtype patients. Thirty-one mRNA transcripts covering 13 immune-metabolic homeostasis genes and 18 immune checkpoint/infiltration-related genes were divided into two functional modules. Spearman correlation analysis was performed to assess co-expression patterns among candidate genes. Receiver operating characteristic (ROC) curves combined with five-fold cross-validation were used to compare the discriminatory efficacy of single-gene markers and the three-gene combined panel.

**Results:** Strong positive co-expression was observed between IRF1, CD8A and LAG3 (IRF1-CD8A: r=0.93; IRF1-LAG3: r=0.84; CD8A-LAG3: r=0.73). Nominal P-values indicated elevated expression of IRF1, CD8A and LAG3 in IE subtype, though no intergroup significance remained after Benjamini-Hochberg FDR correction, largely attributed to the limited sample size of IE cases. Single-gene ROC analysis showed AUC values of 0.763 (IRF1), 0.752 (CD8A) and 0.771 (LAG3), with LAG3 exhibiting the best individual discriminatory capacity. The three-gene combined panel yielded a cross-validated AUC of 0.717, inferior to single LAG3, due to severe collinearity that generated redundant predictive information.

**Conclusions:** The lung cancer-originated TME subtyping system cannot be directly extrapolated to Chinese CRC patients. LAG3 serves as a promising independent transcriptomic candidate marker for distinguishing CRC TME subtypes. The robust collinearity among IRF1, CD8A and LAG3 eliminates additional predictive benefits of the combined signature. Large independent multi-center Chinese CRC cohorts are required to construct population-specific immune transcriptomic biomarkers for standardized clinical NGS TME stratification.

## Main

The incidence and mortality of CRC rank among the top of global malignant tumors ^[1]^, accounting for approximately 10% of the total cancer - related deaths. CRC is no longer regarded merely as an adenomatous epithelial cell malignant proliferative disease driven by gene mutations. Instead, it is a complex TME ecosystem interwoven by tumor cells, immune cells, stromal cells, ECM (Extra Cellular Matrix), and gut microbiota ^[2–3]^. Currently, the TME typing algorithms and gene cut - off values adopted by the mainstream next - generation sequencing (NGS) detection platforms in China are basically based on the pan - cancer TME typing system published in □Cancer - Cell 2021□^[4]^. The main body of the original training set consists of non - small - cell lung cancer (NSCLC) samples from European populations, not established based on Chinese CRC samples. The effectiveness of this typing system when applied to CRC remains unclear. Based on existing domestic and foreign research, this study hopes to horizontally compare the TME subtyping efficiency of lung cancer characteristic genes in CRC and explore exclusive transcriptional markers suitable for CRC TME immune stratification.

## 1 Materials and Methods

### 1.1 Clinical Data

The relevant clinical data of CRC surgical patients in the First Medical Center of the Chinese PLA General Hospital from July 2023 to April 2024 were collected. Finally, 87 cases with complete clinical data, clear pathological diagnosis, and sufficient tumor tissue samples were included. Among them, 55 were male and 32 were female, with an age range of 34 - 84 years and a median age of 61.9 years. NGS testing was carried out. Exclusion criteria^[5]^: (1) Not conforming to the diagnosis of primary adenocarcinoma of CRC; (2) Incomplete clinicopathological data. Tumor tissue paraffin blocks were sectioned into 5 - μm - thick slices, and the tumor proportion in the tissue was required to be ≥20%. The proportion of tumor cells was evaluated by pathologists. The research protocol was approved by the Ethics Committee of Chinese People’s Liberation Army General Hospital (Ethics No.2025080109). All enrolled patients signed written informed consent before NGS detection, and all clinical data were anonymized to protect patient privacy.

### 1.2 Reagents and Instruments

FFPE DNA/RNA automated nucleic acid extraction kit (AmoyDx, Xiamen, China), automatic nucleic acid extractor (AmoyDx, Xiamen, China, model EASY12), pan - solid tumor tissue sample (Master Panel) panoramic gene detection kit (high - throughput sequencing method) (AmoyDx, Xiamen, China), micro - ultraviolet spectrophotometer, real-time fluorescence quantitative PCR instrument (Stratagene Mx3000P™, Agilent Technologies, USA), Illumina gene sequencer (model: Illumina NextSeq CN500), micro - centrifuge, vortex mixer, etc.^[5].^

### 1.3 Next - generation Sequencing Detection and Bioinformatics Analysis

The genomic DNA and RNA of tissue samples were extracted by an automatic nucleic acid extractor. After nucleic acid separation, a nucleic acid quantification kit was used to determine the total amount of DNA and RNA, and the integrity of the nucleic acids was quality - controlled. The total amount of DNA extracted from tissue samples should be ≥60 ng, and the total amount of RNA extracted from tissue samples should be ≥200 ng. Immediately after sample quality control, the experimental operations were carried out approximately in three parts: tissue DNA library construction, tissue RNA library construction, and hybridization capture, following the detection method of the pan - solid tumor tissue sample (Master Panel) panoramic gene detection kit (high - throughput sequencing method). After the experiment, the Illumina NextSeq CN500 sequencing platform was used to sequence the target library ^[5]^. The average sequencing depth of tissue specimens was 500x, the lower limit of detection for the mutation proportion was 5%, and the detected mutation types included single - nucleotide variants (SNV), small - fragment insertions/deletions (Indel), copy - number variations (CNV), gene fusions, etc. The relevant diagnosis and treatment indicators generated by bioinformatics analysis included gene mutation status, microsatellite instability (MSI), tumor microenvironment (TME), tumor mutational burden (TMB), and other mutation types of unknown significance. Based on the limitations of the number of test samples and the principle of clinical practicality, this study simplified and combined the TME results in the NGS test results into two major subtyping groups: the IE subtype and the D+F subtype.

### 1.4 Selection of Research Genes

Based on the existing background and domestic and foreign literature, we selected a total of 31 mRNA transcription indicators, including 13 immune - metabolic regulatory genes and 18 broad - spectrum immune - related genes. These were divided into an immune - metabolic homeostasis module (Module A) and an immune checkpoint - immune infiltration module (Module B), mainly forming two gene sets: □ Genes related to immune activation transcription and metabolic homeostasis (13): This includes the colorectal - cancer - specific immune transcription factor IRF1, the intestinal epithelial marker REG4, the key glycolysis gene ALDOA, and homeostasis housekeeping genes such as PPIA, B2M, ACTB, CD58, GAPDH, HPRT, NOTCH3, UBC, TPM2, and TPM4. These genes are used to analyze the unique metabolic - immune regulatory axis in colorectal cancer. □ Genes related to immune cell infiltration, T - cell exhaustion, and immune checkpoints (18): This includes the core markers of the commercial lung - cancer - derived TME typing system (PDCD1, CD274, CTLA4, CD8A, LAG3, etc.), as well as genes covering T cells, macrophages, chemokines, and multiple immune checkpoint molecules, such as CD4, FOXP3, CD86, CD163, MS4A4A, CD19, NCAM1, CTLA4, HAVCR2, CXCL9, CXCL10, GZMB, IFNG, and TGFB1.

### 1.5 Statistical Analysis

Data analysis was performed using SPSS 25.0 statistical software. For count data, the chi - square test or Fisher’s exact test was used. A P - value less than 0.05 was considered statistically significant. Python 3.10 software was used to draw visual statistical charts. Spearman correlation analysis was employed to analyze the co - expression relationships among the 31 genes. The diagnostic efficacy of the selected genes alone and in combined models in differentiating the IE and D+F subtypes was evaluated through the ROC curve combined with five-fold cross-validation.Shapiro-Wilk test was applied to test the normality of gene expression data; skewed distribution was observed, so non-parametric Mann-Whitney U test was adopted to verify the results of Welch’s t-test.

## 2 Results

### 2.1 Comparison of clinicopathological characteristics among different TME stratifications

We enrolled 87 CRC patients (15 IE-subtype and 72 F+D-subtype cases). We compared histological type, sex, lymph-node metastasis, tumor site, clinical stage, TMB and MSI status between groups using Fisher’s exact test. As shown in Table 1, no parameters differed significantly (all P□ >□0.05). Our results confirm that TME-defined subgroups are independent of traditional clinical confounders. Gene-expression differences, instead of tumor progression, drive varied TME phenotypes in our CRC cohort.

**Tab.1.**
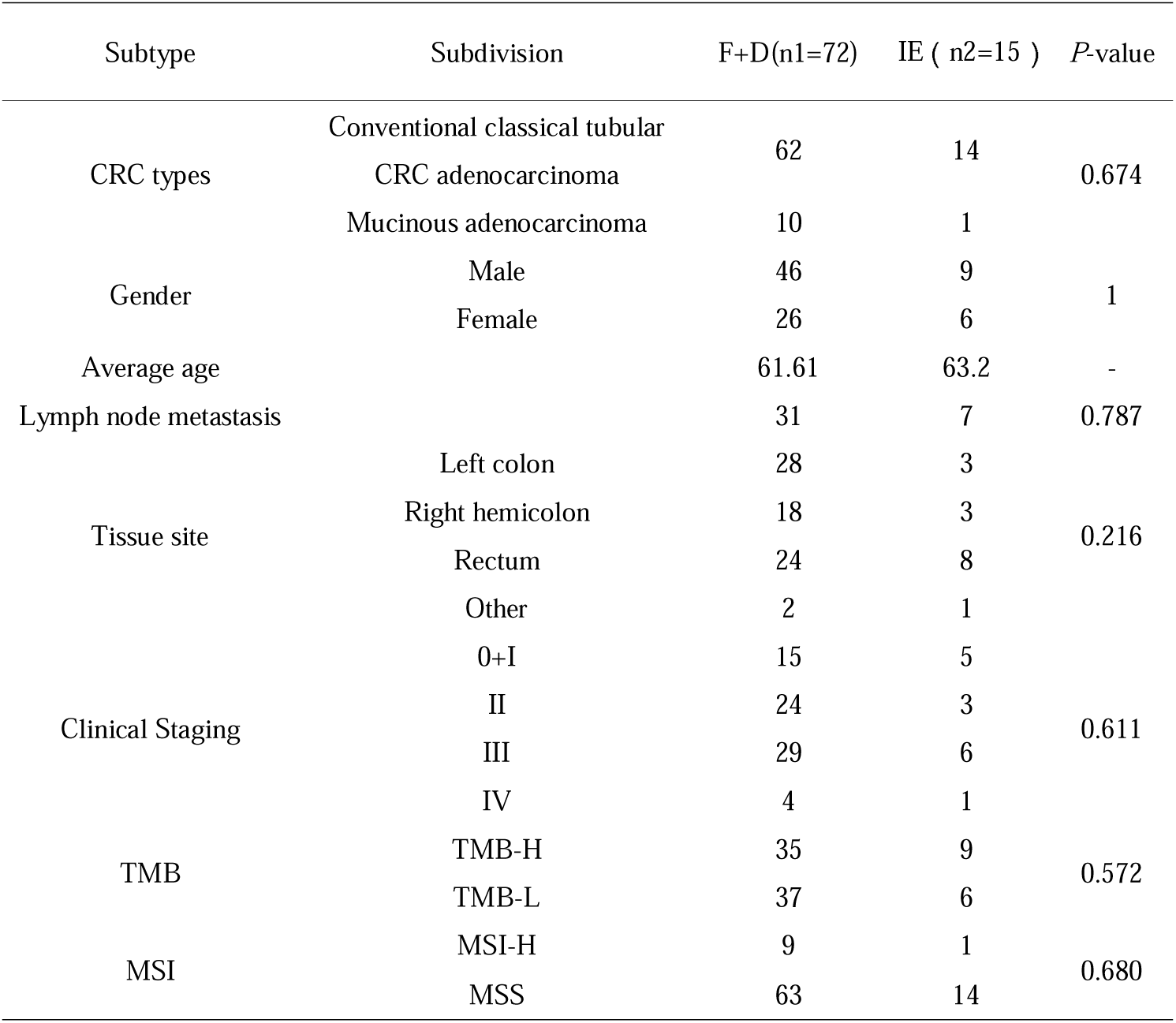
Comparison of clinicopathological characteristics among different TME stratifications.

### 2.2 Comparative Analysis of Filtered Module-Related Genes in Different TME Subgroups

In the present study, the 31 mRNA markers were categorized into Module A containing 13 genes linked to immune-metabolic homeostasis and Module B consisting of 18 genes associated with immune infiltration and immune checkpoints. The Welch-corrected independent-samples t-test was performed to compare gene-expression levels between the two subgroups. The Benjamini-Hochberg FDR approach was used for multiple-test correction to minimize type-I errors. The differential gene-expression profiles between groups are presented in Tables 2 and 3.

**Tab. 2.**
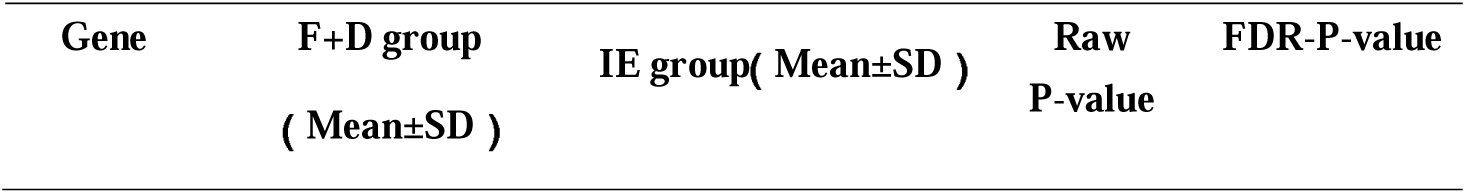
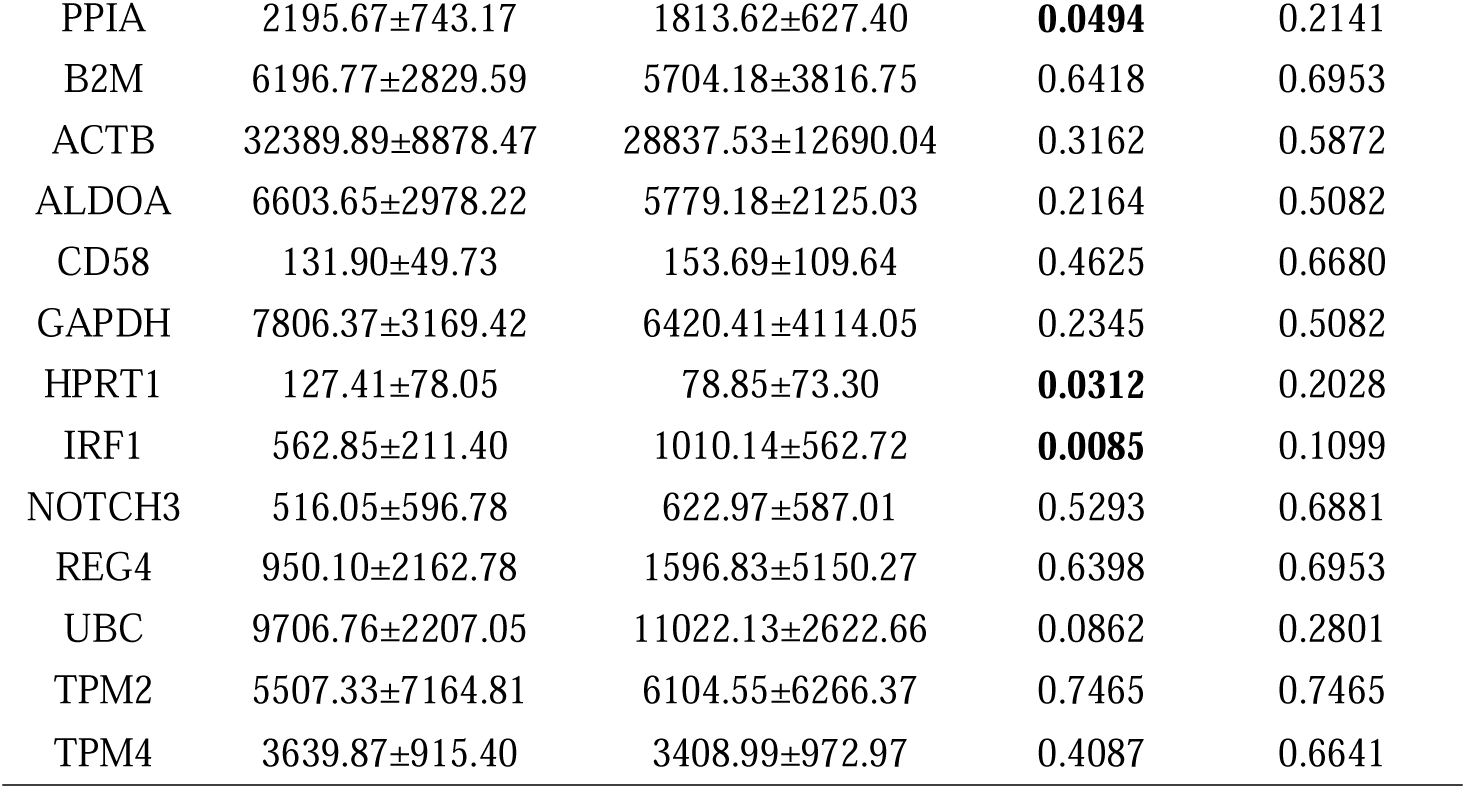
Analysis of Inter-group Expression Differences of Genes in Module A.

**Tab. 3.** Analysis of Inter-group Expression Differences of Genes in Module B.

| Gene | F+D group ( Mean±SD ) | IE group ( Mean±SD | Raw P-value | FDR-P-value |
| --- | --- | --- | --- | --- |
| LAG3 | 19.24±22.61 | 49.33±48.96 | **0.0340** | 0.3653 |
| CD8A | 37.97±65.91 | 84.71±76.53 | **0.0406** | 0.3653 |
| CD4 | 143.5±109.51 | 210.41±124.48 | 0.0686 | 0.4116 |
| CD19 | 20.76±37.73 | 41.83±42.81 | 0.0931 | 0.4192 |
| PDCD1 | 20.09±25.33 | 63.09±113.82 | 0.1664 | 0.5269 |
| CTLA4 | 19.14±14.53 | 27.45±21.77 | 0.1756 | 0.5269 |
| IFNG | 1.74±1.58 | 5.97±13.23 | 0.2361 | 0.5313 |
| CD86 | 37.13±23.82 | 26.73±32.37 | 0.2544 | 0.5313 |
| FOXP3 | 14.31±10.39 | 34.53±67.92 | 0.2691 | 0.5313 |
| HAVCR2 | 44.12±51.48 | 67.81±82.59 | 0.3009 | 0.5313 |
| CD274 | 15.59±19.90 | 29.37±51.60 | 0.3247 | 0.5313 |
| CXCL9 | 175.33±218.42 | 316.27±588.58 | 0.3752 | 0.5628 |
| CD163 | 164.61±105.99 | 202.06±208.74 | 0.5084 | 0.7040 |
| GZMB | 91.29±208.70 | 121.22±199.03 | 0.6050 | 0.7495 |
| TGFB1 | 307.51±146.27 | 356.82±381.20 | 0.6289 | 0.7495 |
| NCAM1 | 52.72±91.57 | 46.57±35.68 | 0.6662 | 0.7495 |
| MS4A4A | 46.12±40.54 | 47.61±41.93 | 0.9009 | 0.9074 |
| CXCL10 | 125.25±126.49 | 132.12±217.67 | 0.9074 | 0.9074 |

In Module A, IRF1 was significantly overexpressed in the IE subgroup relative to the F+D subgroup, whereas PPIA and HPRT1 exhibited lower expression within the IE group. For Module B, CD8A and LAG3 were markedly up-regulated in the IE group with raw P < 0.05. After multiple-testing correction using the Benjamini-Hochberg FDR method, the adjusted P-values of all genes exceeded 0.05, indicating no statistically intergroup differences. Despite the loss of statistical significance following FDR adjustment, elevated expression of IRF1, CD8A and LAG3 in the IE subtype was consistent with its biological feature of immune enrichment. As housekeeping homeostasis-related genes, PPIA and HPRT1 were down-regulated in IE samples, which reflected altered metabolic status upon tumor immune activation and implied distinct expression patterns of immune-metabolic homeostasis genes between these two TME subtypes. We further performed the Mann-Whitney-U test for sensitivity analysis, and the overall expression trends were consistent with those obtained from Welch’s t-test. Given that this was a single-center cohort with merely 15 IE samples, insufficient statistical power was the primary contributor to non-significant outcomes after FDR correction.

We constructed a volcano plot (Figure 1) and scatter plots (Figure 2-6) for the five candidate genes with initial significant differences (IRF1, CD8A, LAG3, PPIA and HPRT1) to visualize mRNA expression levels across individual patient samples.

**Fig. 1.**
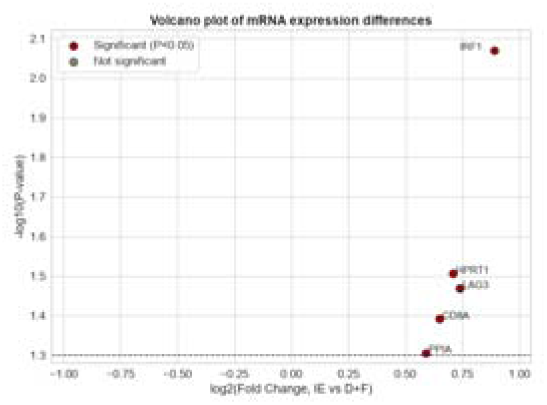
Volcano plot showing gene-expression changes between IE and F+D subgroups.

**Fig. 2.**
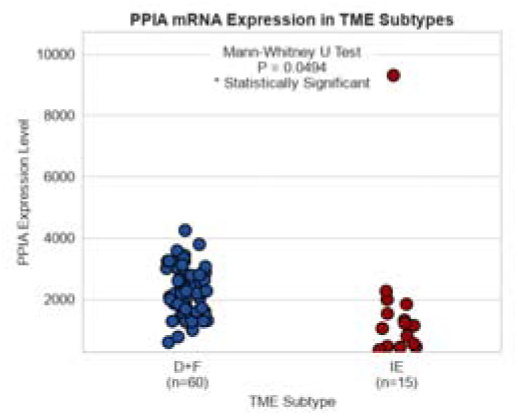
Scatter plot of PPIA gene expression

**Fig. 3.**
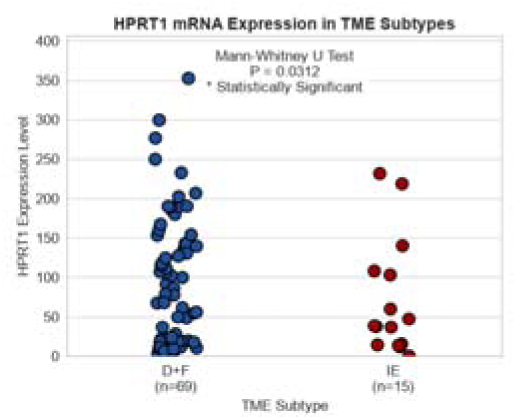
Scatter plot of HPRT1 gene expression

**Fig. 4.**
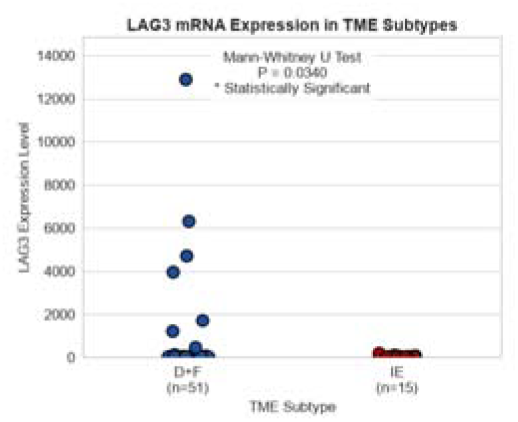
Scatter plot of LAG3 gene expression

**Fig. 5.**
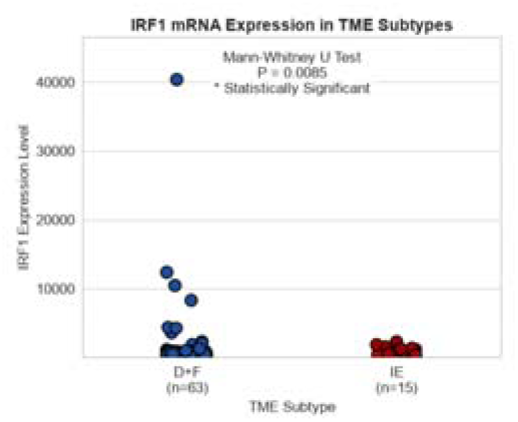
Scatter plot of IRF1 gene expression

**Fig. 6.**
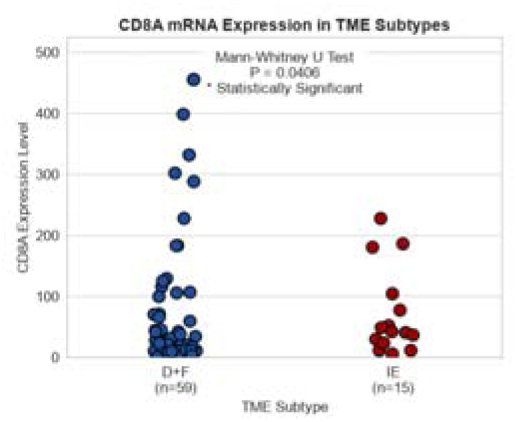
Scatter plot of CD8A gene expression

As shown in Figure 1, we screened differentially-expressed genes across IE and F+D subgroups based on 31 mRNA markers. The x-axis corresponds to log2 fold-change (IE vs F+D), while −log10-transformed P-values from Mann-Whitney-U test are plotted on the y-axis. Genes with raw P < 0.05 are highlighted in red, and IRF1 presents the greatest inter-group discrepancy among these genes.

Noticeable outliers were observed in partial samples from the volcano plot and scatter plots (Figures 1-6), which validated skewed distribution of our expression data and justified adopting non-parametric rank-sum tests instead of t-tests. Only raw P-values from individual-gene comparisons were visualized in these graphs. No genes showed significant differences after FDR correction for the 31 candidates. We finally picked out IRF1, LAG3 and CD8A (excluding house-keeping genes) as key differentially-expressed genes for further analyses.

### 2.3 Further investigation of candidate genes with differential expression across TME subgroups

Spearman correlation analysis was conducted for these 31 genes to investigate inter-gene co-expression patterns.

As illustrated in Figure 7, IRF1, CD8A and LAG3 were strongly and positively correlated (r = 0.93 for IRF1-CD8A, r = 0.84 for IRF1-LAG3 and r = 0.73 for CD8A-LAG3), whereas most other genes had poor pairwise correlations. House-keeping metabolic genes in Module A were moderately correlated with each other but weakly associated with this gene trio. High collinearity suggested concordant expression of IRF1-CD8A-LAG3 in CRC specimens.

**Figure 7.**
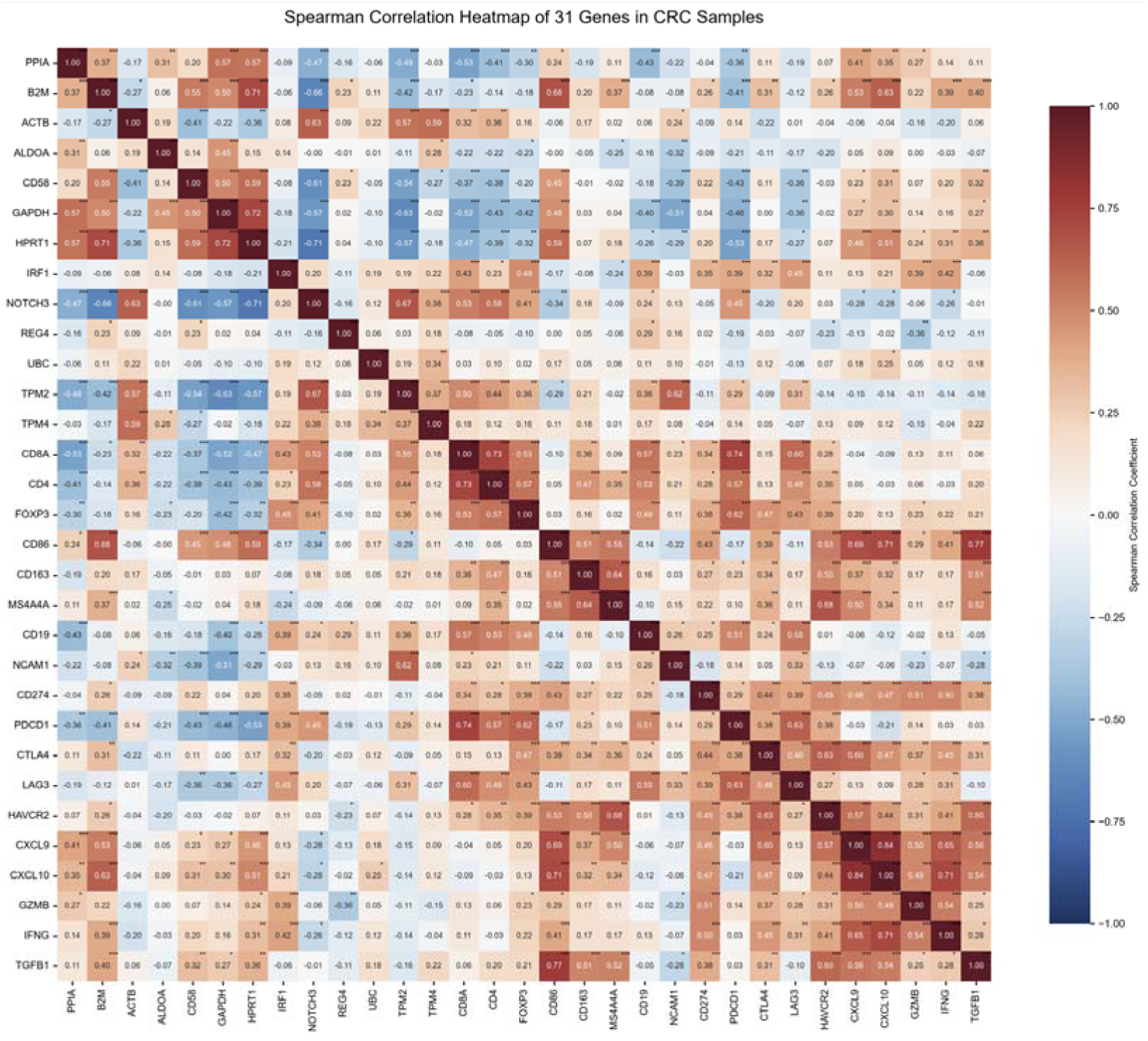
Spearman correlation heatmap of 31 mRNA-related genes. The color gradient represents Spearman correlation coefficient from −1.0 (dark-blue) to 1.0 (dark-red).

After screening out candidate genes with differential expression by volcano plots and confirming their expression distribution via dot-plots, we identified prominent collinearity among IRF1, CD8A and LAG3 from Spearman correlation analysis. On this basis, we performed ROC-curve analysis with five-fold cross-validation to compare the predictive performance of single-gene markers and the three-gene combined panel, and judge whether the lung-cancer-derived gene signature is applicable for Chinese colorectal-cancer patients.

ROC-curve analysis with five-fold cross-validation was carried out to evaluate the diagnostic performance of IRF1, CD8A, LAG3 and their combined signature for distinguishing IE and F+D subtypes. As shown in Figure 8, the AUC values of IRF1, CD8A and LAG3 were 0.763, 0.752 and 0.771 respectively, which indicated that LAG3 achieved the best discriminatory capacity among single-gene markers. Unexpectedly, the AUC of three-gene combined panel decreased to 0.717 after cross-validation and was inferior to single LAG3. Consistent with our Spearman-correlation analysis, high collinearity among IRF1, CD8A and LAG3 led to redundant predictive information, which explained why the lung-cancer-derived three-gene signature failed to improve predictive efficiency in CRC patients. Our results suggested that single LAG3 rather than the combined-gene panel was a promising biomarker for Chinese CRC patients.

**Figure 8.**
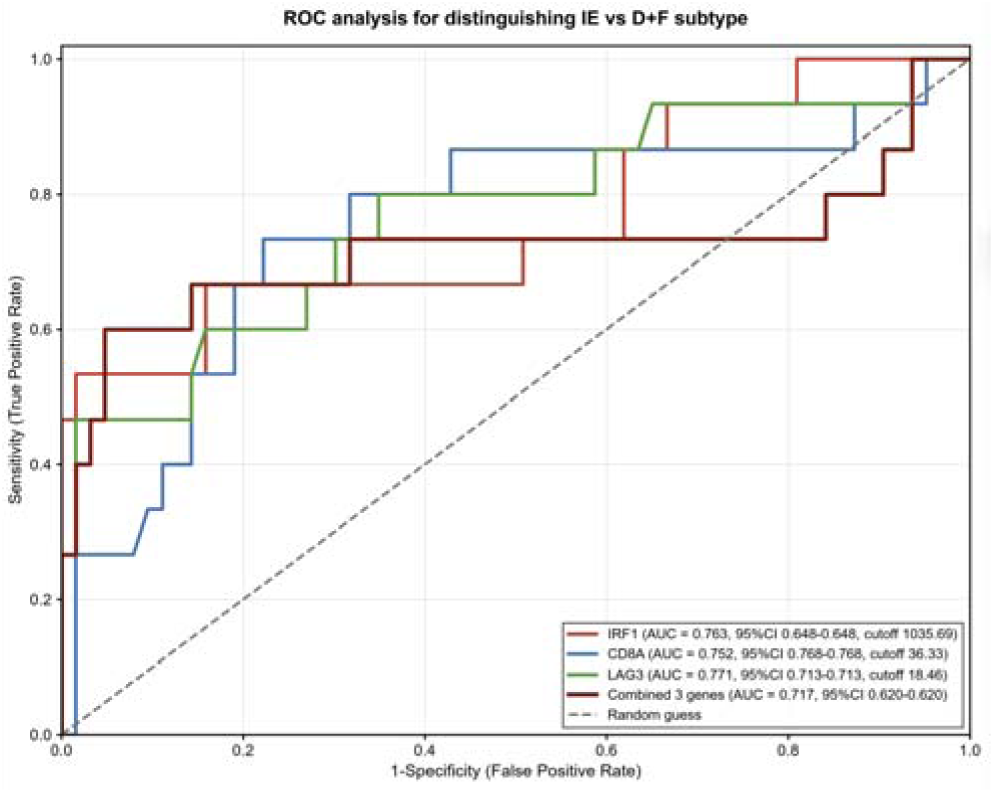
ROC-curve analysis of single-gene markers and three-gene combined signature for differentiating IE and F+D subtypes. Notes : five-fold cross-validation was performed to avoid over-fitting. The dashed line represents random prediction.

### 2.4 Mechanism-related schematic illustration

Based on our mRNA expression profiles and correlation analysis, we herein propose a mechanistic model (illustrated in Figure 9) to delineate the formation of two distinct TME subtypes(the IE subtype and F+D subtype) in Chinese patients with CRC.

**Figure 9.**
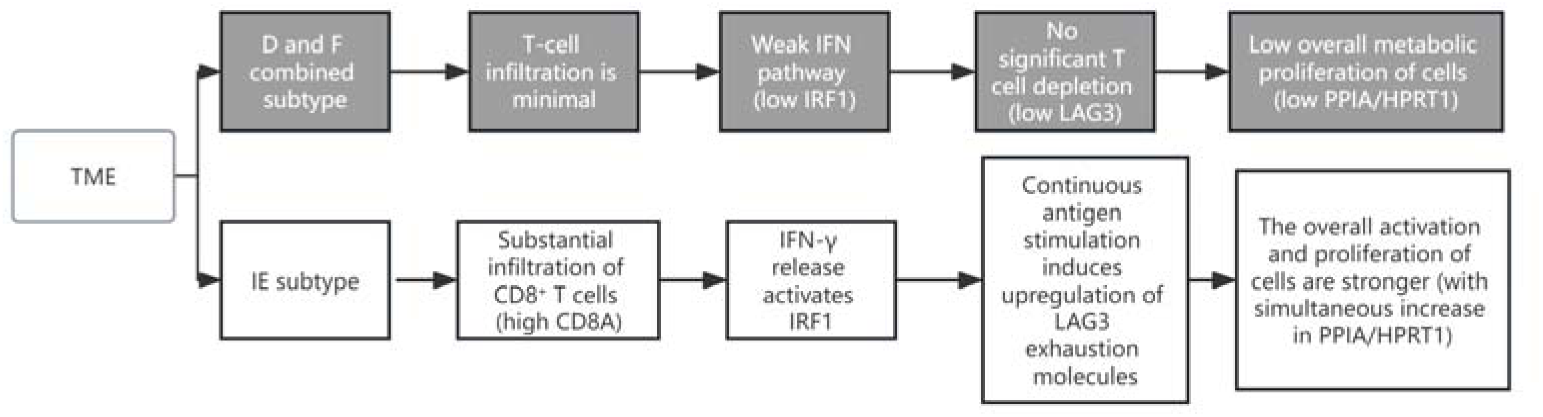
Schematic diagram showing molecular-regulatory pathways underlying IE and F+D TME-subtypes in colorectal-cancer patients.

The regulatory chain from CD8 -T-cell infiltration to IFN-γ-IRF1-LAG3 axis and subsequent metabolic-gene changes was summarized.

In IE-subtype samples, abundant CD8 T-cell infiltration (high CD8A) leads to elevated IFN-γ secretion, which further activates the transcription factor IRF1. Persistent antigen stimulation then up-regulates LAG3 and results in T-cell exhaustion. Meanwhile, activated immune cells increase their proliferation and metabolic activity accompanied by elevated PPIA and HPRT1 expression. In contrast, the F+D subtype shows minimal CD8 T-cell infiltration, which leads to weak IFN-γ-IRF1 pathway activity, low-level LAG3 expression and decreased metabolic-related gene expression. This upstream-downstream regulatory relationship explains why IRF1, CD8A and LAG3 are highly correlated in our CRC cohort, while their correlation is relatively moderate in lung-cancer cohorts reported by previous studies. Tumor-specific differences in IFN-γ-driven signalling contribute to higher collinearity among these three genes in CRC samples, which further accounts for poor performance of the lung-cancer-derived three-gene signature after five-fold cross-validation. Taken together, this model not only explains the molecular basis of IE-F+D stratification, but also supports that single LAG3 rather than three-gene panel is more suitable for Chinese-patient-based TME-classification. Our findings remind domestic NGS platforms that CRC-specific cut-off values should be established instead of directly adopting the standard derived from European lung-cancer patients.

## 3 Discussion

TME plays a vital role in regulating CRC initiation, evolution, metastasis and drug resistance. Recent studies have revealed a multilayer regulatory network for CRC development from failed immune surveillance to the construction of pre-metastatic niches^[2,6].^ However, high spatial-temporal heterogeneity of CRC-TME^[7–8]^, unrealistic simulation of human intestinal flora by experimental models, and varied standardization among detection platforms restrict our ability to fully reproduce dynamic multi-dimensional features of human TME.

In recent years, multiple Chinese studies on CRC-TME in local cohorts have been published. Yet the majority suffer from single-center design and small sample sizes. No consensus-based guideline-level mRNA subtyping standard exclusive to Chinese CRC patients has been formulated domestically, which accounts for the prevailing practice that domestic NGS providers apply European lung-cancer-derived criteria.

In view of these circumstances and published literature, we screened 31 mRNA transcripts (13 immune-metabolic-modulating genes plus 18 pan-immune-related genes) and assigned them to Module A (immune-metabolic homeostasis) and Module B (immune-checkpoint and immune-infiltration module). Our statistical analyses and scatter-plot outcomes revealed inverse expression patterns of immune-related genes (IRF1, CD8A, LAG3) versus metabolic-related genes (PPIA, HPRT1) between TME subgroups, implying metabolic reprogramming occurs alongside immune activation in the CRC-TME.

Prior studies identified IRF1, CD8A and LAG3 as core molecules remodeling CRC-TME^[9–13]^. Being a downstream transcription factor of IFN-γ, IRF1 drives CD8 T-cell recruitment by inducing CXCL9 and CXCL10 transcription to establish immune-enriched TME. CD8A mRNA levels quantify cytotoxic CD8□ T-cell infiltration, and LAG3 marks exhausted CD8□ T cells in CRC. Most previous work investigated these genes individually. The IRF1-CD8A-LAG3 signature was initially developed from European lung-cancer datasets yet directly adopted by domestic commercial NGS platforms for CRC-TME classification without sufficient verification in Chinese patients.

We observed extremely high collinearity among IRF1-CD8A-LAG3 (r = 0.93, 0.84 and 0.73), with stronger co-expression than that in lung-cancer cohorts. Owing to redundant information, the three-gene panel was less predictive than LAG3 alone after five-fold cross-validation, proving that tumor-specific co-expression weakens the utility of lung-cancer-derived markers in CRC. Despite elevated mRNA-expression of IRF1, CD8A and LAG3 in IE-subtype patients, only LAG3 reliably differentiates IE and F+D subtypes in Chinese CRC samples. FDR-BH adjustment across all 31 genes abolished significant differences largely because of the small IE-group sample size (n = 15). Larger Chinese cohorts are warranted to validate our results.

The application of state-of-the-art techniques greatly promotes our understanding of CRC-TME. Combined scRNA-seq and spatial transcriptomics^[14]^ delineate heterogeneous cell subsets and complicated interactive networks within the TME, while multiplex immunofluorescence and mass-cytometry^[15]^ harvest massive microenvironmental data from small biopsies. Still, it is imperative to discover specific TME biomarkers for screening, therapy and prognosis.

World-wide consensus has been achieved on MSI-testing sites and cutoff values. Physicians prescribe pembrolizumab relying on NGS-based MSI-H/MSS status because this classification works across different tumor types. However, the IRF1-CD8A-LAG3 signature presents obvious tumor-specific co-expression features. Lung-cancer-derived markers lose predictive value in CRC. Consequently, we should build CRC-exclusive transcriptomic biomarkers instead of adopting lung-cancer gene sets for NGS-based TME stratification.

Limitations of the present study should be noted. First, this is a single-center retrospective exploratory cohort with a small number of IE subtype patients (n=15), which reduces statistical power and leads to non-significant results after FDR correction. Second, only mRNA transcript levels were analyzed, without protein-level verification via immunohistochemistry or multiplex immunofluorescence. Third, long-term follow-up data on immunotherapy response and patient survival were not collected, so the prognostic value of LAG3 could not be evaluated. Multi-center large-scale prospective cohorts are needed to validate our conclusions.

## Supporting information

Ethical Approval

## Data Availability

All data generated in this study are available from the authors upon reasonable request.

