## Supplementary material for "LAG3 as an independent TME biomarker in Chinese colorectal cancer: Validation of a lung cancer-derived subtyping signature": Ethical Approval

**中国人民解放军总医院 Chinese PLA General Hospital**

**中国人民解放军医学院 Chinese PLA Medical School**

NO. 2025080109

负责人: 中国人民解放军总医院第一医学中心病理科 李金龙

兹证明上述研究项目经过中国人民解放军总医院医学伦理委员会的审核批准,该项目对患者的健康没有危险性，病例收集日期从 2023 年 7 月 1 日至 2024 年 4 月 30 日， 项目（实验）有效期：2025 年 8 月 1 日至 2026 年 8 月 1 日。

中国人民解放军总医院医学伦理委员会

2025 年 8 月 1 日

Project title:

Principal Investigator: Li JinLong（LI JL）, MD
Department of Pathology
First Medical Center, Chinese People's Liberation Army General Hospital

This project was approved by the Medical Research Ethnic Committee of Chinese PLA General Hospital. This project has no risk on patient’s health. The case collection period is from July 1, 2023 to April 30, 2024, and the validity period of the project (experiment) is from August 1, 2025 to August 1, 2026.

The Medical Research Ethnic Committee
Chinese PLA General Hospital
August 1, 2025

**No. 28 Fuxing Road, Haidian District, Beijing 100853, P. R. China**
